# Neutrophil-lymphocyte ratio across psychiatric diagnoses: An electronic health record investigation

**DOI:** 10.1101/2020.01.07.20016790

**Authors:** Aimee Brinn, James M Stone

## Abstract

**Objectives:** The main objective of this study was to compare neutrophil-lymphocyte ratio, a marker of systemic inflammation, between patients diagnosed with ICD-10 psychiatric disorders and control participants.

**Design:** A cross-sectional methodology was employed to retrospectively analyse electronic health records and records derived from a national health survey.

**Setting:** A secondary mental health care service consisting of four boroughs in South London.

**Participants:** A diverse sample of 13,888 psychiatric patients extracted from South London and Maudsley electronic health records database and 3,920 control participants extracted from National Health and Nutrition Survey (2015-2016) were included in the study.

**Primary and secondary outcome measures:** Primary: NLR levels in patients with mental health diagnoses, NLR between patients with different mental health diagnoses. Secondary: Relationship of NLR to length of hospitalisation and to mortality.

**Results:** NLR was elevated compared to controls in patients with diagnoses including dementia, alcohol dependence, schizophrenia, bipolar affective disorder, depression, non-phobic anxiety disorders, and mild mental retardation (p < 0.05). NLR also correlated with age, antipsychotic use and hypnotic use. NLR was found to be higher in individuals of “White” ethnicity and lower in individuals of “Black” ethnicity. Elevated NLR was associated with increased mortality (β = 0.103, p = 2.9e-08) but not with hospital admissions or face-to-face contacts.

**Conclusions:** Elevated NLR may reflect a transdiagnostic pathological process occurring in a subpopulation of psychiatric patients. NLR may be useful to identify and stratify patients who could benefit from adjunctive anti-inflammatory treatment.

**Article Summary:** *Strengths and limitations of this study:*

- Largest study to date of cross-diagnostic neutrophil-lymphocyte measurements in a psychiatric population.
- Sample is representative of diverse adult psychiatric patients in South London.
- Systematic differences between patients and controls reduce the validity of these comparisons.
- This study was retrospective and thus, confounding measures such as body mass index, smoking status and diet were unavailable.

## Introduction

Inflammation is a key component of our immune response to physiological injury or infection (Xiao, 2017). Low-grade systemic inflammation is an attenuated, but persistent, form of the inflammatory response and has been found to be prevalent across a range of psychiatric diagnoses, including psychotic, mood, neurotic, and personality, disorders (Osimo et al, 2018).

Blood biomarkers are the most commonly used method to study inflammatory processes in psychiatry (Fond et al, 2018, Pinto et al, 2017), but many of these are costly or difficult to collect routinely. The neutrophil to lymphocyte ratio (NLR) can be calculated from a full blood count and is thus cheaper and more readily available than cytokine testing (Gibson et al, 2007). NLR has been suggested to be a marker of the intensity of physiological stress present in acutely ill patients, and a measure of systemic inflammation (Zahorec, 2001). As it constitutes parameters from both innate (neutrophil) and adaptive (lymphocyte) immune systems, it may be less affected by confounding variables, such as exercise, compared to other commonly used inflammatory biomarkers (Gibson et al, 2007). NLR has demonstrated reliable prognostic value across a range of physical health disorders (Afari & Bhat, 2016, Chandrashekara et al, 2017, Koh et al, 2015, Wang et al, 2017), highlighting a positive association between systemic inflammation and worse clinical outcomes.

The most studied inflammatory biomarkers in psychiatry, interleukin-6 (IL-6), tumour necrosis factor (TNF) and c-reactive protein (CRP), are elevated in patients with schizophrenia, bipolar disorder and depression, compared to controls (Pinto et al, 2017). NLR also appears to be elevated in major psychiatric disorders compared to controls (Karageorgiou et al, 2018, Mazza et al, 2018, Mazza et al, 2019a). However, the vast proportion of studies have studied a single disorder in comparison to healthy controls, or at best, two different psychiatric disorders (Pinto et al, 2017). True transdiagnostic studies are rare with only three studies to date comparing NLR between psychiatric diagnoses (Baykan et al, 2018, Mazza et al, 2019b, Ozdin et al, 2017).

Of the NLR studies comparing different psychiatric diagnoses, Ozdin et al (2017) reported that patients with schizophrenia, experiencing a psychotic episode, had higher NLR than patients with manic-phase bipolar disorder (BD), suggesting there are detectable differences in NLR across diagnosis and symptomatology. Mazza et al (2019b) found patients with manic-phase BD had higher NLR than patients with major depressive disorder (MDD). They also found significant differences between manic-phase and depressive-phase BD but not between depressive-phase BD and MDD suggesting that NLR may be elevated in mania but not depression. Lastly, Baykan et al (2018) found NLR to be significantly elevated in elderly patients with Alzheimer’s (AD), compared to elderly patients with MDD. Shared symptomatology between AD and MDD is problematic for clinicians differentiating between the disorders (Baykan et al, 2018) and the authors suggested that NLR may be an effective diagnostic marker to aid discrimination.

The current evidence base is limited and inconsistent. Further research is required to investigate the role of inflammatory processes in psychiatric illness. Differentially elevated inflammation between different diagnoses may suggest inflammatory processes are more common in specific disorders. Alternatively, comparable elevation between diagnoses may suggest common aetiological pathways, such as shared immunological genetic vulnerabilities and inflammatory processes. Patients with elevated NLR may represent a subtype of psychiatric illness that does not fit within current diagnostic categories. Lastly, systemic inflammation may be a by-product of mental health difficulties, reflecting a non-specific comorbid physiological stress.

In this study, we aimed to characterise NLR across a range of ICD-10 (World Health Organisation, 1992) psychiatric diagnoses and to investigate the relationship between NLR and adverse clinical outcomes.

## Methods

### Design

This study employed a cross-sectional methodology to analyse electronic health records extracted using the Clinical Record Interactive Search (CRIS) tool (Stewart et al, 2009). We retrospectively analysed neutrophil and lymphocyte counts of psychiatric patients using whole blood count data. For comparison, a control group were obtained from the National Health and Nutrition Examination Survey (Centers for Disease Control and Prevention: National Center for Health Statistics, 2016).

### Clinical Record Interactive Search

The Clinical Record Interactive Search (CRIS) application was developed in 2007, with funding from the British National Institute for Health Research. The application generates a data resource consisting of anonymised electronic health records of service users of the SLaM NHS Foundation Trust (Stewart et al, 2009). These records are contained in the Biomedical Research Centre (BRC) case register, which is updated every 24 hours. CRIS was used to access patient data, using SQL management studio as a platform for data extraction.

CRIS has ethical approval for usage as an anonymised database for secondary analysis (reference 18/SC/0372). This project was approved by the CRIS Oversight Committee.

### Patient Sample

The SLaM BRC case register includes all patients who have accessed secondary mental health care services within a catchment area of four south London boroughs (Lambeth, Southwark, Lewisham and Croydon). SLaM is one of the largest mental healthcare organisations in Europe, providing care to 1.3 million people (Lopez-Morinigo et al, 2018).

We extracted data from records for all patients who accessed SLaM secondary mental health care between 01-01-2007, when CRIS was established, and 20-11-2018, when our data were extracted.

Data were extracted for all patients meeting the following inclusion criteria:

- ≥ 16 years old at earliest recorded blood test
- ICD-10 Primary Diagnosis F01-F79
- Recorded neutrophil count
- Recorded lymphocyte count

13,888 patients met our inclusion criteria and were included in the sample.

### Control Sample

The control group comprised a sample of participants from the 2015–2016 National Health and Nutrition Survey (Centers for Disease Control and Prevention: National Center for Health Statistics, 2016). The original sample included 9,971 participants from across 30 locations in the United States. Hispanic, Black and Asian individuals are overrepresented in the sample, as are individuals at or below 185 percent of the Department of Health and Human Services poverty guidelines (Burwell, 2015) and those aged 80 and over.

We included individuals meeting the following inclusion criteria:

- ≥16 years old at time of NHANES (2015-2016) survey
- Recorded neutrophil count
- Recorded lymphocyte count

Office for National Statistics (ONS) (Office for National Statistics, 2009) guidelines do not code ‘Hispanic’ as a distinct ethnic group. ‘Hispanic’ was therefore not a distinct ethnic group in the CRIS database. For ease of sample comparison, we excluded Hispanic participants from our study. 3,920 participants met our criteria and were included in the control sample.

### Variables

#### Demographics

Demographic variables were retrieved from structured fields in the CRIS database and included age at earliest blood test, gender and ethnicity. Ethnicity was coded using ONS guidelines (Office for National Statistics, 2009). To facilitate comparison with the NHANES sample, we combined ‘Other’ and ‘Mixed’ ethnicity codes into one category ‘Other/Mixed’.

#### Neutrophil to lymphocyte ratio

Absolute neutrophil and lymphocyte counts were obtained from routine blood testing data, attained at the point of patient entry to SLaM secondary care services. NLR was calculated for each patient by dividing neutrophil count by lymphocyte count. We extracted earliest available blood count values to minimise potential confounding effects of secondary healthcare treatment.

#### ICD-10 diagnosis

We included diagnoses from eight ICD-10 diagnostic blocks in this study: Organic, including symptomatic, mental disorders (F01-F09), Mental and behavioural disorders due to psychoactive substance use (F10-F19), Schizophrenia, schizotypal and delusional disorders (F20-F29), mood [affective] disorders (F30-F39), Neurotic, stress-related and somatoform disorders (F40-F49), Behavioural syndromes associated with physiological disturbances and physical factors (F50-59), Disorders of adult personality and behaviour (F60-F69) and Mental retardation (F70-F79). Diagnostic variables were defined by the primary diagnosis assigned to the patient closest to their blood test result.

We excluded patients with diagnoses in the Disorders of psychological development (F80-F89) and Behavioural/emotional disorders with onset usually occurring in childhood and adolescence (F90-F98) diagnostic blocks as most patients were under 16 years old. We also excluded patients in the Unspecified mental disorder (F99) block due to ambiguity of the diagnosis.

#### Adverse clinical outcomes

We defined ‘number of overnight bed stays’ as the total number of overnight hospital stays for each patient since the date of the blood test. We defined ‘number of face-to-face events’ as the number of clinical face-to-face contacts, such as hospital appointments, for each patient since the date of the blood test. Both measures were adjusted for patient time in services since the date of the blood test. Death was recorded as a dichotomous mortality variable.

#### Medication

The medication prescribed to the patient within 1 year of the blood test date was extracted. Medications were categorised into four classes: antipsychotics, antidepressants, mood stabilisers and hypnotics.

#### Statistical Analysis

We conducted all statistical analyses for this study using R (version 3.6). We analysed data distribution by use of histogram. We report the mean and standard deviation (SD) for normally distributed data.

Because NLR data demonstrated an exponential distribution (see Figures 1 and 2),we conducted regression analyses using a log-linked gamma-family generalised linear model. For each analysis, the shape parameter of the model was estimated using the gamma.shape function from the MASS package, and this was used to set the dispersion parameter as 1/(maximum likelihood estimate). We ran the following models: 1) Control NLR as dependent variable vs. NLR in ICD-10 diagnoses; 2) NLR between all ICD-10 diagnoses that were significantly different from control NLR from Model 1. In Models 1 and 2 age, ethnicity, gender and medication were included as independent variables. We then ran three separate models within the CRIS dataset only to investigate the association of NLR with clinical outcome measures. These models set NLR as the dependent variable vs death, hospital admissions or numbers of face-to face contact. ICD-10 diagnosis, age, ethnicity, gender and medication were included as independent variables.

**Figure 1:**
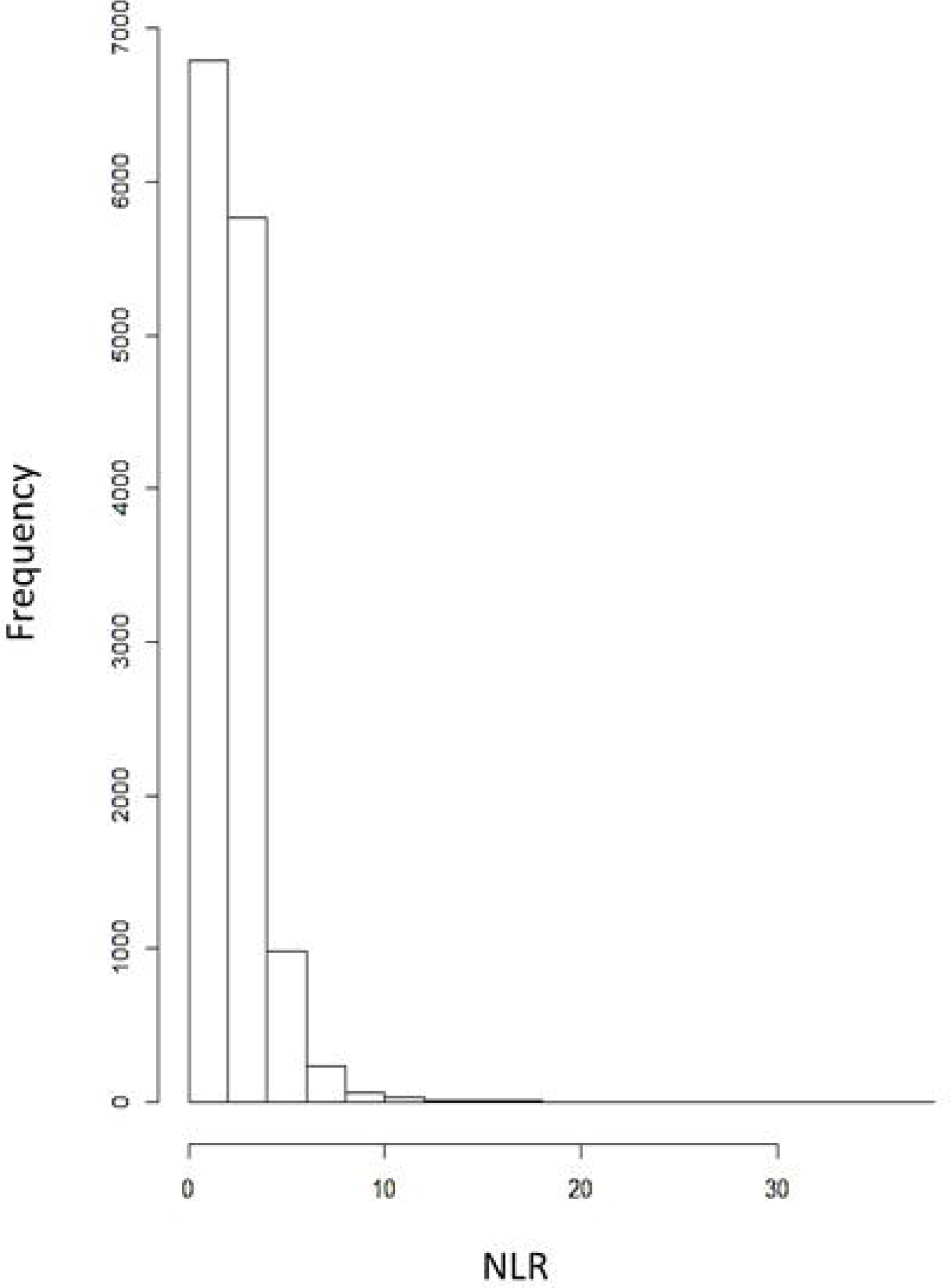
Distribution of Neutrophil to Lymphocyte Ratio (NLR) in patient sample

**Figure 2:**
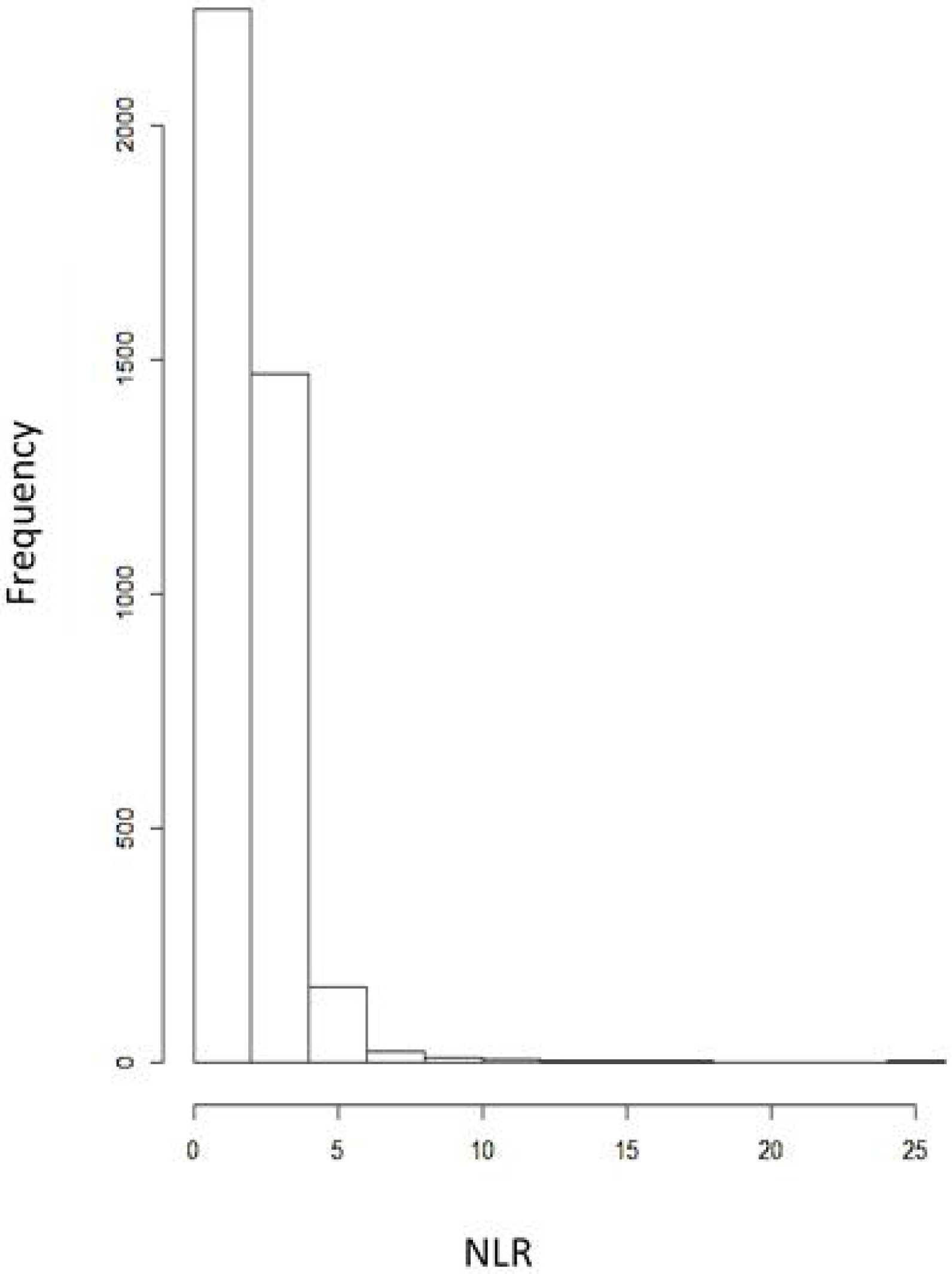
Distribution of Neutrophil to Lymphocyte Ratio (NLR) in control sample

## Results

### Demographic Results

The mean ages of patients and controls were 40.1 (SD: 16.57) and 46.9 (SD: 19.47) respectively. Distribution of demographic variables are displayed in Table 1.

**Table 1.** Distribution of age, ethnicity and gender variables across patient and control samples. (N = number in group, % = relative percentage of total sample).

|  | Patient Sample |  | Control Sample |  |
| --- | --- | --- | --- | --- |
|  | N | % | N | % |
| Age Group |  |  |  |  |
| 16-19 | 1201 | 8.65 | 350 | 8.93 |
| 20-29 | 3188 | 22.95 | 600 | 15.31 |
| 30-39 | 2989 | 21.52 | 591 | 15.08 |
| 40-49 | 2759 | 19.87 | 619 | 15.79 |
| 50-59 | 1987 | 14.31 | 584 | 14.90 |
| 60-69 | 878 | 6.32 | 537 | 13.70 |
| 70-79 | 561 | 4.04 | 366 | 9.34 |
| 80+ | 325 | 2.34 | 273 | 6.96 |
| Ethnicity |  |  |  |  |
| Asian | 788 | 5.67 | 668 | 17.04 |
| Black | 3749 | 26.99 | 1187 | 30.28 |
| Other/mixed | 1238 | 8.91 | 217 | 5.54 |
| White | 8113 | 58.43 | 1848 | 47.17 |
| Gender |  |  |  |  |
| Male | 6539 | 47.08 | 1945 | 49.6 |
| Female | 7346 | 52.9 | 1975 | 50.4 |
| Non-specified | 3 | 0.02 | - | - |

There was no relationship between date of blood test (from which the NLR value was derived) and NLR value across the patient sample (β = −4.76e-06, p = 0.692) suggesting that NLR measurement remained stable across the defined time period (2007-2018).

### Model 1: NLR between controls and patients

NLR in the following ICD-10 diagnoses were significantly higher than controls: F00, Alzheimer’s disease (β = 0.15, p = 1.3e-06); F01, vascular dementia (β = 0.24, p = 0.003); F02 dementia in other diseases (β = 0.27, p = 0.008); F05, delirium (β =0.28, p = 0.003); F06, other mental disorders due to brain damage and dysfunction (β = 0.16, p = 0.0006); F10, mental and behavioural disorders due to alcohol (β = 0.16, p = 2.0e-14); F20, schizophrenia (β = 0.09, p = 9.7e-08); F23, acute and transient psychotic disorders (β = 0.16, p = 3.7e-08); F25, schizoaffective disorders (β = 0.07, p = 0.002); F28, other nonorganic psychotic disorders (β = 0.17, p = 0.03); F29, unspecified nonorganic psychotic disorders (β = 0.13, p = 1.9e-07); F30, manic episode (β = 0.20, p = 3.0e-06); F31, bipolar affective disorder (β = 0.12, p = 3.0e-07); F32, depressive episode (β = 0.13, p =7.31e-11); F33, recurrent depressive disorder (0.14, p = 7.9e-07); F38 other mood [affective] disorders (β = 0.41, p = 0.001); F41, other (non-phobic) anxiety disorders (β = 0.06, p =0.03); F43, reaction to severe stress, and adjustment disorders (β = 0.08, p = 0.004); F60, specific personality disorders (β = 0.09, p = 0.0005); F61, mixed personality disorders (β = 0.18, p = 0.03); F70, mild mental retardation (β = 0.14, p = 0.02) and F79, unspecified mental retardation (β = 0.40, p = 0.047). There were no diagnoses with NLR significantly lower than controls. In the model, age was also positively correlated with NLR (β = 0.005, p < 2e-16; Figure 3). Black ethnicity was associated with lower NLR (β = −0.23, p < 2e-16) and White ethnicity with higher NLR (β = 0.09, p = 5.8e-07; Figure 4). Antipsychotic use (β = 0.04, p = 0.003)) and hypnotic use (β = 0.03, p = 0.01) were both associated with higher NLR (see supplementary materials table T1).

**Figure 3:**
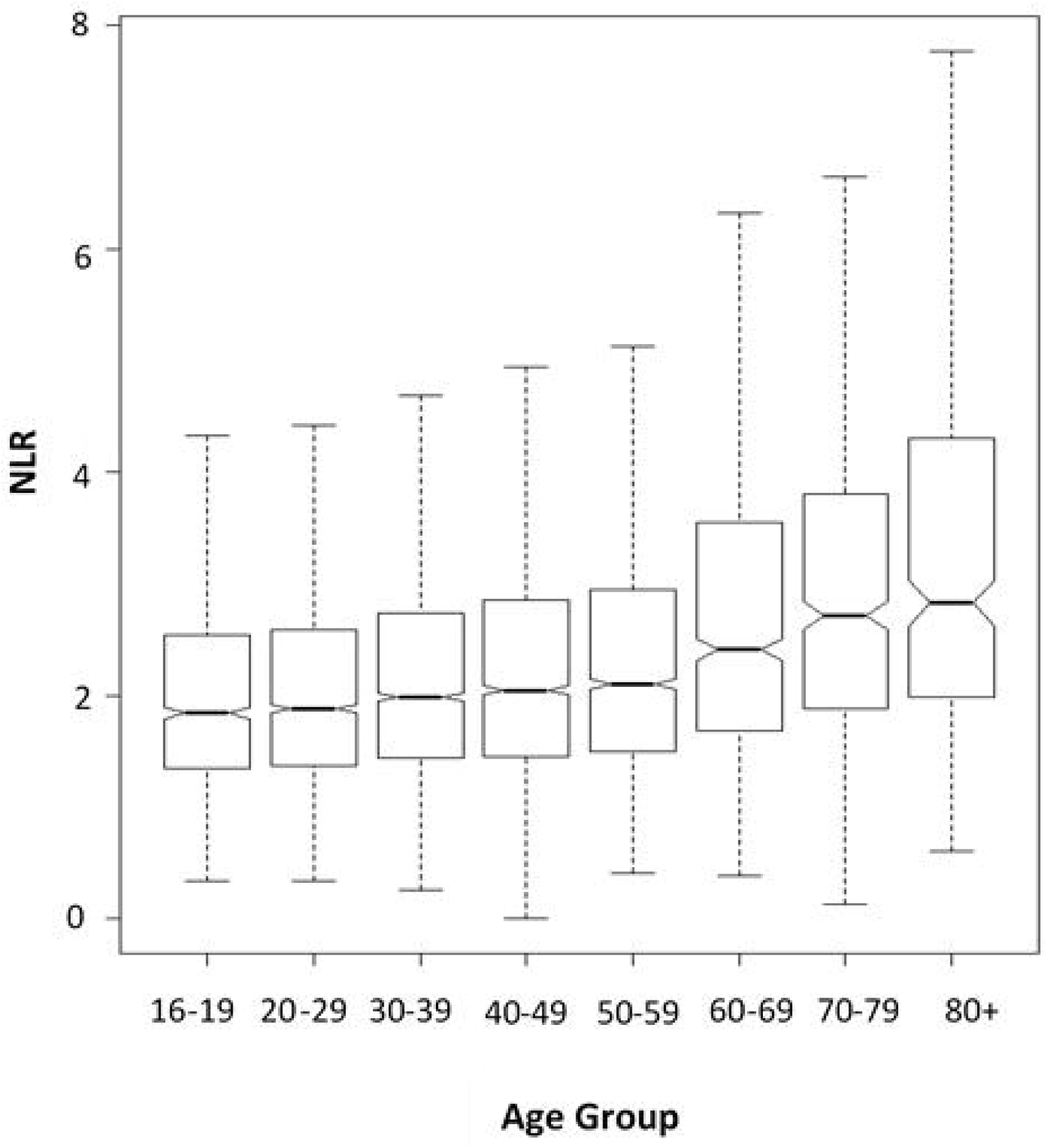
Neutrophil to Lymphocyte Ratio (NLR) (uncorrected) across ethnicity

**Figure 4:**
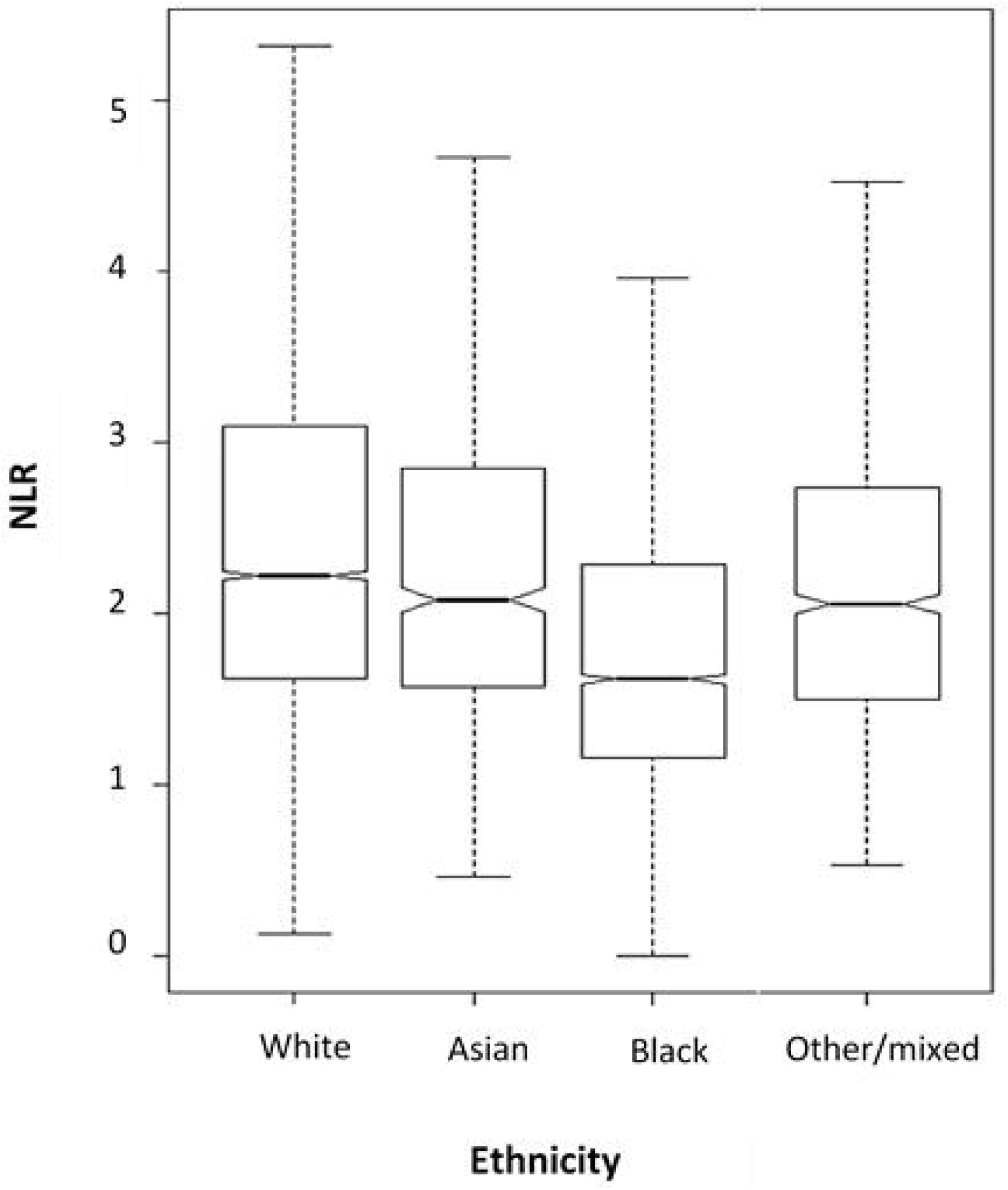
Neutrophil to Lymphocyte Ratio (NLR) (uncorrected) across age groups

### Model 2: NLR between diagnoses with elevated NLR

We found that of the diagnoses elevated in Model 1, patients with a diagnosis of F38, other mood [affective] disorders, had higher NLR (β = 0.27, p = 0.04), whereas all other diagnoses did not differ significantly (see supplementary materials table T2).

### Model 3: NLR across adverse clinical outcomes

There was a positive association between mortality and NLR, adjusting for ICD-10 diagnostic group, age, gender, ethnicity, antipsychotic, antidepressant, mood stabiliser and hypnotic medication use, all of which contributed to the model according to AIC score (β = 0.103, p = 2.9e-08; see supplementary materials table T3).

There was no significant relationship between number of overnight bed stays or number of face-to-face events and NLR.

## Discussion

This study suggests that systemic inflammation occurs in several psychiatric diagnoses. In particular, NLR elevation was seen in patients with dementia, alcohol dependence, schizophrenia, bipolar affective disorder, depression, non-phobic anxiety disorders, and mild mental retardation. In those groups with elevated NLR, the only diagnostic group that had significantly higher NLR was F38 “other mood disorders”. All other diagnoses with elevated NLR did not significantly differ in terms of NLR level, suggesting that NLR elevation is a non-specific, cross-diagnostic finding. It is possible that patients with elevated NLR have a shared inflammatory aetiology to their different clinical presentations.

In comparing our findings to published data, one previous study of NLR found that patients with acute schizophrenia had higher NLR than patients with manic phase BD (Ozdin et al, 2017), whereas Mazza and colleagues (2019b) did not find any difference in NLR between patients with bipolar and unipolar depression. Another group reported that NLR was elevated in patients with Alzheimer’s disease compared to age-matched patients with MDD and Parkinson’s disease (Baykan et al. 2018). We did not find any evidence for elevated NLR in Alzheimer’s disease compared to other diagnoses, but we were not able to exclude the possibility that NLR elevations may occur during acute exacerbations of psychosis.

Our results align closely with findings from other inflammatory biomarkers. A recent meta-analysis indicated variation patterns in inflammatory biomarkers IL-6, TNF and CRP were comparable across diagnoses (Pinto et al, 2017). Thus, current evidence suggests that phenomenologically defined diagnostic criteria do not map closely onto biologically-based pathogenic markers such as NLR (Zachar & Kendler, 2017).

In terms of the association of NLR with patient outcomes, we found a significant positive relationship been NLR and mortality but we found no relationship between NLR and ‘number of overnight bed stays’ or ‘number of face-to-face events’. A previous study investigated a similar clinical outcome measure ‘number of hospitalisations’ and found a positive relationship with NLR in patients with BD (Melo et al, 2019). The study involved a small sample of 80 patients, but the findings were strengthened by the fact the authors employed a longitudinal design. The difference in findings with our study could be due to the cross-diagnostic nature of our study, and the fact that the patients were not all in-patients.

A positive relationship between mortality and NLR has been indicated across physical health disorders (Afari & Bhat, 2016, Koh et al, 2015, Templeton et al, 2014). As we were not able to adjust for physical health comorbidities, the relationship that we found may reflect an association between systemic inflammation and poor physical health. Alternatively, our findings may reflect an independent positive relationship between NLR and mortality in psychiatric patients. In both BD patients and MDD patients, NLR has been found to be a significant predictor of suicide risk (Ekinci & Ekinci, 2017, Ivkovic et al, 2016), which may account for some of the increased mortality. However, elevated NLR has been found to independently predict mortality, irrespective of age, sex, education, metastatic cancer, liver disease and depression (Isaac et al, 2016) suggesting systemic inflammation may reduce survival through alternative pathological pathways.

## Limitations

This study has several limitations. Confounding factors which may modulate NLR such as body mass index (Azab et al, 2014), smoking (Tulgar et al, 2016), diet and exercise (Gialluisi et al, 2019) were not controlled for and so interactions such as a positive correlation between depression and obesity (Luppino et al, 2010, Mannan et al, 2016) may have confounded our results. Likewise, comorbid physical conditions found to modulate NLR, such as cancer or autoimmune disease (Bhat et al, 2013, Celikbilek et al, 2013, Templeton et al, 2014), were neither excluded nor adjusted for.

The control sample represents a non-institutionalised civilian (US) population. These participants were not screened for physical or mental disorders, so the prevalence of these conditions in the control sample is unknown. The fact that samples were selected from different countries and blood cell essays were performed in different laboratories reduces the validity of our patient and control comparisons. However, although NLR was elevated in some ICD-10 diagnoses compared to the control group, NLR in patients with other diagnoses did not significantly differ from this control sample, suggesting that the control NLR was suitable as a comparison group in this study.

This study did not employ validated symptom severity measures. ‘Number of overnight bed stays’ and ‘number of face-to-face events’ are logical outcome measures of illness severity. These real-world measures increased the clinical transferability of our results, but they may be affected by unknown factors, such as help-seeking attitudes or illness insight.

This sample is representative of a population seeking secondary care services. Results may not be generalisable to patients seeking primary care services. Finally, this is a cross-sectional study, so causal inferences cannot be inferred.

Nonetheless, this is the largest study to date of cross-diagnostic NLR measurements in a psychiatric population and our sample is representative of adult psychiatric patients in South London. Every patient who accessed SLaM services within our time frame and had available blood count data from age 16 years old or above, was included in our sample. This minimised selection biases. Previous studies included only elderly (Baykan et al, 2018) or Caucasian patients (Mazza et al, 2019b) and so were less generalisable. Our sample is diverse in age and ethnicity so demonstrates improved translational potential. This study also compares NLR across the broadest range of diagnoses to date and employs the largest sample yet to address these two research aims.

## Future Directions

Findings suggest broad transdiagnostic commonalities regarding systemic inflammation. As within-diagnostic differences in inflammation have also been found (Raison & Miller, 2011, Lamers et al, 2013), it appears inflammatory processes are not bound to traditional diagnostic categories. In both depression and schizophrenia, inflammation is thought to contribute to treatment resistance (Mondelli et al, 2015, Lanquillion, Krieg, Bening-Abu-Shach & Vedder, 2000), perhaps contributing to poor primary treatment response rates in psychiatry (Gaynes et al, 2008, Tiihonen et al, 2017).

It may be fruitful for future studies investigating NLR in psychiatric populations to employ a Research Domain Criteria (RDoC) approach (Cuthbert & Insel, 2010). RDoC reconceptualises psychopathology into transdiagnostic domains, presenting a novel classification of mental illnesses, based on dimensions of observable behaviour and neurobiological measures (Morris & Cuthbert, 2012). As our results indicate inflammatory processes are shared across diagnoses, inflammation may play a role in symptom domains shared by these conditions. For example, alterations in serotonergic, dopaminergic and glutamatergic pathways are consistently reported across mood (Ashok et al, 2017, Cowen, 2008, Werner et al, 2010) and psychotic (Guillin et al, 2007, Stone et al, 2007) disorders and have been reported to be mediated by immunological pathways (Miller, 2009, Miller et al, 2013, Müller & Schwarz, 2007). Immune-mediated alterations in serotoninergic and glutamatergic systems may result in mood and cognitive symptoms whilst inflammation-induced modifications of dopaminergic systems may result in motivational changes (Capuron & Castanon, 2016).

NLR may be a useful and cost-effective transdiagnostic parameter by which to stratify psychiatric patients on a common pathophysiological basis. Recent years have seen pharmaceutical companies withdraw investments from psychiatry due to ‘low probability of success’ (Miller, 2010) and nosology has been highlighted as one of the barriers to research progress (Fibiger et al, 2012). Transdiagnostic biological abnormalities may constitute common targets for pharmacological treatment and NLR may be an accessible tool to aid in identification and stratification of these patients in clinical trials.

## Conclusions

Systemic inflammation may occur as a transdiagnostic pathological process in a subpopulation of psychiatric patients. NLR may be an effective biomarker to identify these patients who may benefit from adjunctive anti-inflammatory pharmacological treatment.

## Data Availability

No additional data is available

## Author Statement

The study was conceived by JS with further development by AB. Data extraction was by AB and was specified by JS. Literature review, data analysis first draft of the manuscript was by AB with supervision by JS. Both authors contributed to manuscript preparation and approved the final version.

## References

Afari, M. E., & Bhat, T. (2016). Neutrophil to lymphocyte ratio (NLR) and cardiovascular diseases: an update. Expert review of cardiovascular Therapy, 14(5), 573–577.

Ashok, A. H., Marques, T. R., Jauhar, S., Nour, M. M., Goodwin, G. M., Young, A. H., & Howes, O. D. (2017). The dopamine hypothesis of bipolar affective disorder: the state of the art and implications for treatment. Molecular psychiatry, 22(5), 666.

Azab, B., Camacho-Rivera, M., & Taioli, E. (2014). Average values and racial differences of neutrophil lymphocyte ratio among a nationally representative sample of United States subjects. PloS one, 9(11), e112361.

Baykan, H., Baykan, Ö., Esen, E. C., Tirak, A., Görgülü, S. A., & Karlıdere, T. (2018). Neutrophil-to-lymphocyte ratio as a potential differential diagnostic marker for alzheimer’s disease, major depressive disorder, and parkinson’s disease. The Journal of Psychiatry and Neurological Sciences. 31. 10.5350/DAJPN2018310407.

Bhat, T., Teli, S., Rijal, J., Bhat, H., Raza, M., Khoueiry, G., & Costantino, T. (2013). Neutrophil to lymphocyte ratio and cardiovascular diseases: a review. Expert review of cardiovascular therapy, 11(1), 55–59.

Burwell, SM. (2015). Annual update of the HHS poverty guidelines. Fed Regist, 80, 3236–3237.

Çakur, U., Can Tuman, T., & Yıldırım, O. (2015). Increased neutrophil/lymphoctye ratio in patients with bipolar disorder: a preliminary study. Psychiatria Danubina, 27(2), 0–184.

Capuron, L., & Castanon, N. (2016). Role of inflammation in the development of neuropsychiatric symptom domains: evidence and mechanisms. In Inflammation-Associated Depression: Evidence, Mechanisms and Implications (pp 31–44). Springer, Cham.

Celikbilek, M., Dogan, S., Ozbakır, O., Zararsız, G., Kücük, H., Gürsoy, S., & Yücesoy, M. (2013). Neutrophil–lymphocyte ratio as a predictor of disease severity in ulcerative colitis. Journal of clinical laboratory analysis, 27(1), 72–76.

Centers for Disease Control and Prevention: National Center for Health Statistics (2017). National Health and Nutrition Examination Survey 2015-2016. (https://wwwn.cdc.gov/nchs/nhanes/ContinuousNhanes/Default.aspx?BeginYear=2015). Accessed June 2019.

Chandrashekara, S., Mukhtar Ahmad, M., Renuka, P., Anupama, K. R., & Renuka, K. (2017). Characterization of neutrophil-to-lymphocyte ratio as a measure of inflammation in rheumatoid arthritis. International journal of rheumatic diseases, 20(10), 1457–1467.

Cowen, PJ. (2008). Serotonin and depression: pathophysiological mechanism or marketing myth?. Trends in Pharmacological Sciences, 29(9), 433–436.

Cuthbert, B. N., & Insel, T. R. (2010). Toward new approaches to psychotic disorders: the NIMH Research Domain Criteria project.

Ekinci, O., & Ekinci, A. (2017). The connections among suicidal behavior, lipid profile and low-grade inflammation in patients with major depressive disorder: a specific relationship with the neutrophil-to-lymphocyte ratio. Nordic journal of psychiatry, 71(8), 574–580.

Fond, G. B., & Boyer, L. (2018). C-reactive protein as a peripheral biomarker in schizophrenia. An updated systematic review. Frontiers in psychiatry, 9, 392.

Fibiger, HC. (2012). Psychiatry, the pharmaceutical industry, and the road to better therapeutics.

Gaynes, B. N., Rush, A. J., Trivedi, M. H., Wisniewski, S. R., Spencer, D., & Fava, M. (2008). The STAR* D study: treating depression in the real world. Cleveland Clinic journal of medicine, 75(1), 57–66.

Gialluisi, A., Bonaccio, M., Di Castelnuovo, A., Costanzo, S., De Curtis, A., Sarchiapone, M., & Moli-Sani Study Investigators. (2019). Lifestyle and biological factors influence the relationship between mental health and low-grade inflammation. Brain, behavior, and immunity.

Gibson, P. H., Croal, B. L., Cuthbertson, B. H., Small, G. R., Ifezulike, A. I., Gibson, G., & Hillis, G. S. (2007). Preoperative neutrophil-lymphocyte ratio and outcome from coronary artery bypass grafting. American heart journal, 154(5), 995–1002.

Guillin, O., Abi-Dargham, A., & Laruelle, M. (2007). Neurobiology of dopamine in schizophrenia. International review of neurobiology, 78, 1–39.

Isaac, V., Wu, C. Y., Huang, C. T., Baune, B. T., Tseng, C. L., & McLachlan, C. S. (2016). Elevated neutrophil to lymphocyte ratio predicts mortality in medical inpatients with multiple chronic conditions. Medicine, 95(23).

Ivković, M., Pantović-Stefanović, M., Dunjić-Kostić, B., Jurišić, V., Lačković, M., Totić-Poznanović, S., Jovanović, A. & Damjanović, A. (2016). Neutrophil-to-lymphocyte ratio predicting suicide risk in euthymic patients with bipolar disorder: Moderatory effect of family history. Comprehensive psychiatry, 66, 87–95.

Karageorgiou, V., Milas, G. P., & Michopoulos, I. (2018). Neutrophil-to-lymphocyte ratio in schizophrenia: A systematic review and meta-analysis. Schizophrenia research.

Koh, C. H., Bhoo-Pathy, N., Ng, K. L., Jabir, R. S., Tan, G. H., See, M. H., … & Taib, N. A. (2015). Utility of pre-treatment neutrophil–lymphocyte ratio and platelet–lymphocyte ratio as prognostic factors in breast cancer. British journal of cancer, 113(1), 150.

Lamers, F., Vogelzangs, N., Merikangas, K. R., De Jonge, P., Beekman, A. T. F., & Penninx, B. W. J. H. (2013). Evidence for a differential role of HPA-axis function, inflammation and metabolic syndrome in melancholic versus atypical depression. Molecular psychiatry, 18(6), 692.

Lanquillon, S., Krieg, J. C., Bening-Abu-Shach, U., & Vedder, H. (2000). Cytokine production and treatment response in major depressive disorder. Neuropsychopharmacology, 22(4), 370–379.

Lopez-Morinigo, J. D., Fernandes, A. C., Shetty, H., Ayesa-Arriola, R., Bari, A., Stewart, R., & Dutta, R. (2018). Can risk assessment predict suicide in secondary mental healthcare? Findings from the South London and Maudsley NHS Foundation Trust Biomedical Research Centre (SLaM BRC) Case Register. Social psychiatry and psychiatric epidemiology, 53(11), 1161–1171.

Luppino, F. S., de Wit, L. M., Bouvy, P. F., Stijnen, T., Cuijpers, P., Penninx, B. W., & Zitman, F. G. (2010). Overweight, obesity, and depression: a systematic review and meta-analysis of longitudinal studies. Archives of general psychiatry, 67(3), 220–229.

Mannan, M., Mamun, A., Doi, S., & Clavarino, A. (2016). Is there a bi-directional relationship between depression and obesity among adult men and women? Systematic review and bias-adjusted meta analysis. Asian journal of psychiatry, 21, 51–66.

Mazza, M. G., Lucchi, S., Tringali, A. G. M., Rossetti, A., Botti, E. R., & Clerici, M. (2018). Neutrophil/lymphocyte ratio and platelet/lymphocyte ratio in mood disorders: A meta-analysis. Progress in Neuro-Psychopharmacology and Biological Psychiatry, 84, 229–236.

Mazza, M. G., Lucchi, S., Rossetti, A., & Clerici, M. (2019a). Neutrophil-lymphocyte ratio, monocyte-lymphocyte ratio and platelet-lymphocyte ratio in non-affective psychosis: A meta-analysis and systematic review. The World Journal of Biological Psychiatry, 1-13.

Mazza, M. G., Tringali, A. G. M., Rossetti, A., Botti, R. E., & Clerici, M. (2019b). Cross-sectional study of neutrophil-lymphocyte, platelet-lymphocyte and monocyte-lymphocyte ratios in mood disorders. General hospital psychiatry, 58, 7–12.

Melo, M. C. A., Garcia, R. F., de Araújo, C. F. C., Abreu, R. L. C., de Bruin, P. F. C., & de Bruin, V. M. S. (2019). Clinical significance of neutrophil-lymphocyte and platelet-lymphocyte ratios in bipolar patients: An 18-month prospective study. Psychiatry research, 271, 8–14.

Miller, AH. (2009). Mechanisms of cytokine-induced behavioral changes: Psychoneuroimmunology at the translational interface. Brain, behavior, and immunity, 23(2), 149–158.

Miller, G. (2010). Is pharma running out of brainy ideas?.

Miller, A. H., Haroon, E., Raison, C. L., & Felger, J. C. (2013). Cytokine targets in the brain: impact on neurotransmitters and neurocircuits. Depression and anxiety, 30(4), 297–306.

Mondelli, V., Ciufolini, S., Belvederi Murri, M., Bonaccorso, S., Di Forti, M., Giordano, A., & Pariante, C. M. (2015). Cortisol and inflammatory biomarkers predict poor treatment response in first episode psychosis. Schizophrenia bulletin, 41(5), 1162–1170.

Morris, S. E., & Cuthbert, B. N. (2012). Research Domain Criteria: cognitive systems, neural circuits, and dimensions of behavior. Dialogues in clinical neuroscience, 14(1), 29.

Müller, N., & Schwarz, M. J. (2007). The immune-mediated alteration of serotonin and glutamate: towards an integrated view of depression. Molecular psychiatry, 12(11), 988.

Office for National Statistics. (2009) Ethnic group, national identity and religion. (https://www.ons.gov.uk/methodology/classificationsandstandards/measuringequality/ethnicgroupnationalidentityandreligion). Accessed April 2019.

Osimo, E. F., Cardinal, R. N., Jones, P. B., & Khandaker, G. M. (2018). Prevalence and correlates of low-grade systemic inflammation in adult psychiatric inpatients: An electronic health record-based study. Psychoneuroendocrinology, 91, 226–234. doi:10.1016/j.psyneuen.2018.02.031

Özdin, S., Sarisoy, G., & Böke, Ö. (2017). A comparison of the neutrophil-lymphocyte, platelet-lymphocyte and monocyte-lymphocyte ratios in schizophrenia and bipolar disorder patients–a retrospective file review. Nordic journal of psychiatry, 71(7), 509–512.

Pinto, J. V., Moulin, T. C., & Amaral, O. B. (2017). On the transdiagnostic nature of peripheral biomarkers in major psychiatric disorders: a systematic review. Neuroscience & Biobehavioral Reviews, 83, 97–108.

Raison, C. L., & Miller, A. H. (2011). Is depression an inflammatory disorder?. Current psychiatry reports, 13(6), 467–475.

Stewart, R., Soremekun, M., Perera, G., Broadbent, M., Callard, F., Denis, M., & Lovestone, S. (2009). The South London and Maudsley NHS foundation trust biomedical research centre (SLAM BRC) case register: development and descriptive data. BMC psychiatry, 9(1), 51.

Stone, J. M., Morrison, P. D., & Pilowsky, L. S. (2007). Glutamate and dopamine dysregulation in schizophrenia—a synthesis and selective review. Journal of psychopharmacology, 21(4), 440–452.

Templeton, A. J., McNamara, M. G., Šeruga, B., Vera-Badillo, F. E., Aneja, P., Ocaña, A., & Tannock, I. F. (2014). Prognostic role of neutrophil-to-lymphocyte ratio in solid tumors: a systematic review and meta-analysis. JNCI: Journal of the National Cancer Institute, 106(6).

Tiihonen, J., Mittendorfer-Rutz, E., Majak, M., Mehtälä, J., Hoti, F., Jedenius, E., & Taipale, H. (2017). Real-world effectiveness of antipsychotic treatments in a nationwide cohort of 29 823 patients with schizophrenia. JAMA psychiatry, 74(7), 686–693.

Tulgar, Y. K., Cakar, S., Tulgar, S., Dalkilic, O., Cakiroglu, B., & Uyanik, B. S. (2016). The effect of smoking on neutrophil/lymphocyte and platelet/lymphocyte ratio and platelet indices: a retrospective study. Eur Rev Med Pharmacol Sci, 20(14), 3112–3118.

Wang, Y., Fuentes, H. E., Attar, B. M., Jaiswal, P., & Demetria, M. (2017). Evaluation of the prognostic value of neutrophil to lymphocyte ratio in patients with hypertriglyceridemia-induced acute pancreatitis. Pancreatology, 17(6), 893–897.

Werner, F. M., & Covenas, R. (2010). Classical neurotransmitters and neuropeptides involved in major depression: a review. International Journal of Neuroscience, 120(7), 455–470.

World Health Organization. (1992). The ICD-10 classification of mental and behavioural disorders: clinical descriptions and diagnostic guidelines. Geneva: World Health Organization.

Xiao, TS. (2017). Innate immunity and inflammation. Cellular & molecular immunology, 14(1), 1–3. doi:10.1038/cmi.2016.45

Zachar, P., & Kendler, KS. (2017). The philosophy of nosology. Annual Review of Clinical Psychology, 13, 49–71.

Zahorec, R. (2001). Ratio of neutrophil to lymphocyte counts-rapid and simple parameter of systemic inflammation and stress in critically ill. Bratislavske lekarske listy, 102(1), 5–14.

